# Machine Learning Prediction of Parkinson’s Disease Onset and Subtype Using Germline Variants

**DOI:** 10.1101/2021.06.14.21258631

**Authors:** Saya R Dennis, Tanya Simuni, Yuan Luo

## Abstract

Parkinson’s Disease is the second most common neurodegenerative disorder in the United States, and is characterized by a largely irreversible worsening of motor and non-motor symptoms as the disease progresses. A prominent characteristic of the disease is its high heterogeneity in manifestation as well as the progression rate. For sporadic Parkinson’s Disease, which comprises 90% of all diagnoses, the relationship between the patient genome and disease onset or progression subtype remains largely elusive. Machine learning algorithms are increasingly adopted to study the genomics of diseases due to their ability to capture patterns within the vast feature space of the human genome that might be contributing to the phenotype of interest. In our study, we develop two machine learning models that predict the onset as well as the progression subtype of Parkinson’s Disease based on subjects’ germline mutations. Our best models achieved an ROC of 0.77 and 0.61 for disease onset and subtype prediction, respectively. To the best of our knowledge, our models present state-of-the-art prediction performances of PD onset and subtype solely based on the subjects’ germline variants. The genes with high importance in our best-performing models were enriched for several canonical pathways related to signaling, immune system, and protein modifications, all of which have been previously associated with PD symptoms or pathogenesis. These high-importance gene sets provide us with promising candidate genes for future biomedical and clinical research.

## 1 Background

Parkinson’s Disease (PD) is the second most common neurodegenerative disease next to Alzheimer’s Disease in the United States, with an occurrence of 160 cases per 100,000 in adults age 65 years or older. [1] The main symptoms of PD can be categorized into motor symptoms, including resting tremor, bradykinesia, and rigidity, and non-motor symptoms, including neuropsychiatric decline, sleep decline, and sensory decline. [2] These symptoms are exacerbated as the disease progresses in a largely irreversible manner. There is no cure for PD to date, and the only available treatments are for relieving the symptoms. Since these treatments are more effective when begun at an earlier stage of the disease, [3] an early assessment of risk for PD is critical.

A person’s genomic profile, typically represented as a list of germline variants, is often rich with information regarding their disease risk, and offers a promising path for risk assessment of PD. A recent increase in publicly available genomic datasets from large-scale PD cohorts has resulted in many studies that provide useful insights on the genomic risk factors as well as the disease etiology of PD. [4, 5] However, the currently known genomic risk factors only explain a small fraction of the disease risk, often referred to as “missing heritability”. [6] This poses a need for better risk models. There are several approaches to improve on existing models, including the incorporation of previously unused features and adoption of a higher-complexity model. Recently, Bandres-Ciga *et al*. achieved the former by performing a systematic, bias-free approach with polygenic-risk-scoring and transcriptomic network analysis to identify certain canonical pathways as risk factors of PD in a large-scale cohort. [4] This work provides us with highly interpretable and actionable results. One limitation of such polygenic risk modeling approach, however, is that it generally does not capture nonlinear relationships between features.

Another prominent research question in the field of PD is its heterogeneity in the type of manifestation and progression rate among patients. [7, 8] Although there have been many attempts at characterizing distinct progression subtypes of PD, [9, 10] a gold standard is yet to be established. A method for assessing a patient’s progression subtype at an early stage of the disease will help clinicians make a more informed decision about a patient’s treatment plan.

Machine learning (ML) has recently been increasingly adopted for studying the genomics of diseases such as cancer, owing to its ability to capture intricate relationships between the numerous components of the genome. [11, 12] The method is particularly advantageous in that it can effectively model nonlinear relationships that might not have been captured by traditional risk modeling approaches. An accurate model to predict either the onset or the progression subtype of PD would prove valuable not only in its predictive capability, but also in providing potential insight into disease etiology by highlighting features that are predictive of the outcome of interest.

Many existing ML studies in PD utilize known clinical, demographic, and genetic risk factors as their model input. [13, 14] Although such models often have satisfactory performance in small-scale cohort studies, this approach fails to explain the “missing heritability” as it cannot find patterns in unknown or uncharacterized features that might be contributing to disease pattern. Therefore, when modeling PD risk based on genomic features, it is crucial to fully utilize whole-exome sequencing (WES) or whole-genome sequencing (WGS) data to develop a model with the best possible performance.

In this study, we utilize WES and WGS data from large-scale cohort studies to train machine learning models for two tasks: PD disease onset prediction and PD progression subtype prediction. We then interpret our best performing models to derive insights that are of clinical and biomedical relevance. To the best of our knowledge, our work presents the state-of-the-art predictive performance of PD onset and progression subtpye solely based on the patients’ germline genomic data.

## 2 Results

### 2.1 Data Description

#### 2.1.1 Acquisition Source

We obtained our data from the Accelerating Medicines Partnership: Parkinson’s Disease (AMP-PD) integrated database. AMP-PD is a harmonized collection of multiple cohort studies of PD including the Michael J. Fox Foundation (MJFF) and National Institutes of Neurological Disorders and Stroke (NINDS) BioFIND study, the NINDS Parkinson’s disease Biomarkers Program (PDBP), and the MJFF Parkinson’s Progression Markers Initiative (PPMI). The database includes genomic sequences, longitudinal clinical test results, and biospecimen data from PD patients as well as healthy controls from multiple, multi-institute studies.

#### 2.1.2 Target Labels

##### Disease Onset Prediction Target

The AMP-PD database provides binary labels for PD patients (cases) and healthy individuals (controls). We discarded observations from subjects whose case/control label changes overtime for simplicity. We utilized WGS data for this task as it was the type of sequencing data most widely available in this cohort. Among the AMP-PD subjects that had germline sequencing data available, the disease onset label was available for 2800 subjects. We refer to this group of subjects as our *Onset Cohort*.

##### Progression Subtype Prediction Target

As there are no agreed-upon progression subtypes for PD, we adopted the progression subtypes characterized in Zhang *et al*.. [13] This study used a deep learning algorithm, Long-Short Term Memory, to represent each patient’s disease progression as a multi-dimensional time-series data from motor and non-motor clinical exams commonly used to measure the severity of PD. This representation was then used to categorize the progression patterns into three distinct subtypes with different manifestations as well as progression rate. The patients’ genomic data was not used in this study. Subtype I was characterized by moderate functional decline in motor ability but a relatively mild cognitive decay, while Subtype II was characterized by mild functional decay in both motor and non-motor abilities, and Subtype III was characterized by a rapid progression in both motor and non-motor symptoms. We utilized WES data for this task as this was the type of sequencing data most widely available in subjects with subtype labels. This cohort consisted of 351 patients, all of whom belong to the PPMI cohort study. We will refer to this group of subjects as the *Subtype Cohort*.

#### 2.1.3 Input Data

Our input data comprises of the number of likely-disruptive variants per gene for each subject. (See Methods for a detailed description of the processing approach.) The variant count distributions of our two cohorts are summarized in Fig. 1. Since WGS sequences a larger proportion of the genome compared to WES, this is likely the reason why the Onset Cohort appears to have a generally higher number of variants compared to the Subtype Cohort. This disparity is not an issue in our study as we train entirely separate models for our two cohorts.

**Figure 1:**
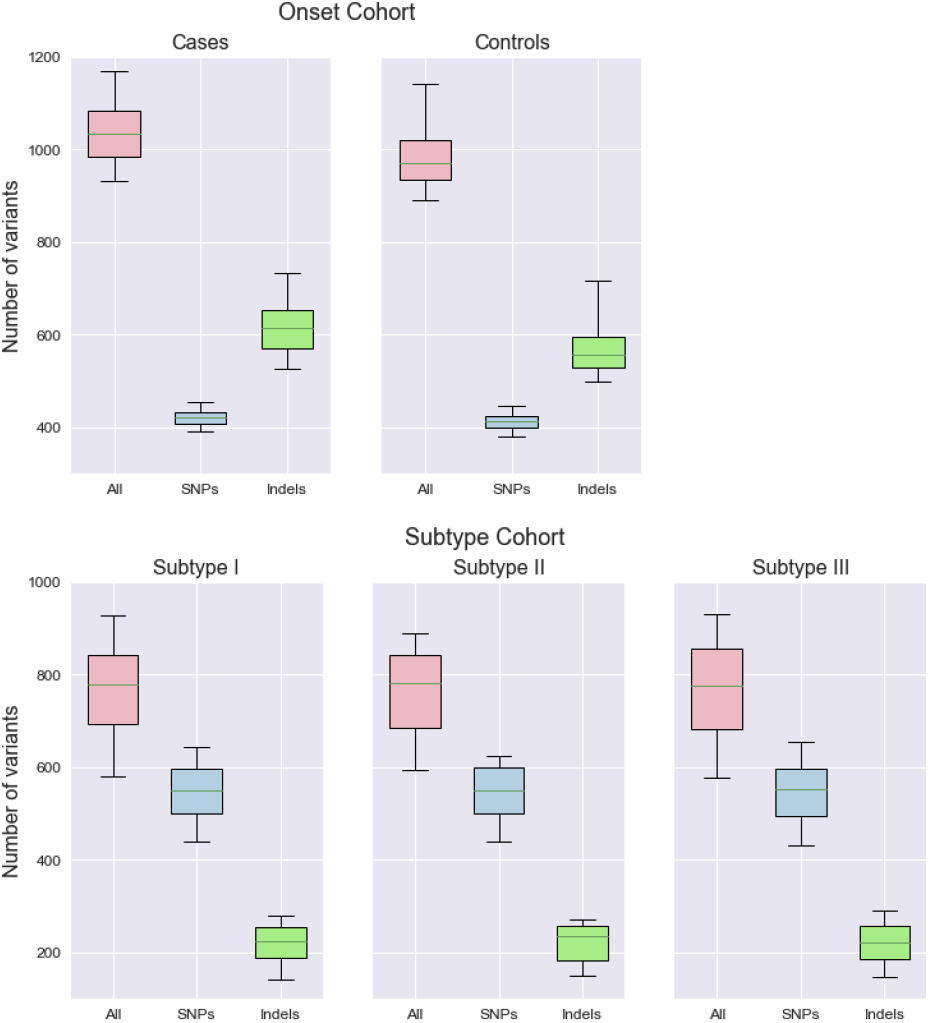
Boxplots showing the number of variants for the two cohorts, separated by case/control or progression subtype. The whiskers extend to the 5% and 95% percentiles.

### 2.2 Cohort Summary

The case-control composition of our two cohorts are summarized in Table. 1. The demographic information of our two cohorts are summarized in Table. 2. For both our Onset Cohort and Subtype Cohort, the age is recorded at the beginning of the longitudinal study.

**Table 1:** Cohort composition summary

| *Onset Cohort* |  |  |
| --- | --- | --- |
| Set | Cases | Controls |
| Overall | 1713 (61%) | 1089 (39%) |
| Train | 1370 (61%) | 871 (39%) |
| Test | 343 (61%) | 218 (39%) |

| *Subtype Cohort* |  |  |  |
| --- | --- | --- | --- |
| Set | Subtype I | Subtype II | Subtype III |
| Overall | 172 (49%) | 47 (13%) | 132 (38%) |
| Train | 151 (48%) | 43 (14%) | 121 (38%) |
| Test | 21 (58%) | 4 (11%) | 11 (31%) |

**Table 2:** Demographic Summary of Cohorts

|  | *Onset Cohort* |  |  | *Subtype Cohort* |  |  |  |
| --- | --- | --- | --- | --- | --- | --- | --- |
|  | Overall | Cases | Controls | Overall | Subtype I | Subtype II | Subtype III |
| **Age at baseline** |  |  |  |  |  |  |  |
| Mean | 62.38 | 63.90 | 59.99 | 61.56 | 58.27 | 63.04 | 65.33 |
| SD | 10.79 | 9.64 | 12.01 | 9.7 | 9.5 | 9.7 | 8.4 |
| **Sex (%)** |  |  |  |  |  |  |  |
| Female | 1249 (44.6) | 661 (38.6) | 588 (54.0) | 102 (29.1) | 52 (14.8) | 17 (4.8) | 33 (9.4) |
| Male | 1553 (55.4) | 1052 (61.4) | 501 (46.0) | 229 (65.2) | 107 (30.4) | 27 (7.7) | 95 (27.1) |
| Unspecified | 0 (0.0) | 0 (0.0) | 0 (0.0) | 20 (5.7) | 13 (3.7) | 3 (0.9) | 4 (1.1) |
| **Race (%)** |  |  |  |  |  |  |  |
| Asian | 25 (0.9) | 22 (1.3) | 3 (0.3) | 6 (1.7) | 2 (1.2) | 0 (0.0) | 4 (3.0) |
| Black | 48 (1.7) | 21 (1.2) | 27 (2.5) | 6 (1.7) | 1 (0.6) | 1 (2.1) | 4 (3.0) |
| Native American | 4 (0.1) | 2 (0.1) | 2 (0.2) | 0 (0.0) | 0 (0.0) | 0 (0.0) | 0 (0.0) |
| Pacific Islander | 1 (0.04) | 0 (0.0) | 1 (0.1) | 0 (0.0) | 0 (0.0) | 0 (0.0) | 0 (0.0) |
| White | 2631 (93.9) | 1606 (93.8) | 1025 (94.1) | 326 (92.9) | 162 (94.2) | 43 (91.5) | 121 (91.7) |
| Multiracial | 58 (2.1) | 34 (2.0) | 24 (2.2) | 0 (0.0) | 0 (0.0) | 0 (0.0) | 0 (0.0) |
| Unknown | 35 (1.3) | 28 (1.6) | 7 (0.6) | 13 (3.7) | 7 (4.1) | 3 (6.4) | 3 (2.3) |

#### Onset Cohort

For our Onset Cohort, there was a statistically significant difference in age between the case and control groups (two-sample Student’s t-test p=5e-21). This is likely due to the fact that, since age is a strong risk factor in PD, older subjects are recruited for the case group. Since our predictor variables are germline mutations and are not affected by age, this difference should not confound our models. The gender composition in the case/control groups also showed a statistically significant imbalance (Fisher’s exact test p=1.5e-15). The larger proportion of male subjects is somewhat representative of the overall diagnosed gender ratio of PD. [15] The class imbalance is corrected for during model training by inversely weighting the observations with their class counts in the loss function.

#### Subtype Cohort

We observed statistically significant differences in age between our subtype groups (one-way ANOVA p=2.6e-10), although it is important to note that subject age was taken into consideration when defining the three subtypes in Zhang *et al*.. Gender shows a roughly consistent 2:1 ratio in all subtypes, which is a sharper skew than overall gender ratio in the diagnoses of PD. [15]

### 2.3 Model Performance

The classification performance of our trained models is summarized in Table. 3. The metrics all indicate performane on the held-out test set, and is macro-averaged for the multiclass classification in the Subtype Cohort. For both the disease onset prediction task and the subtype prediction task, Random Forest and Support Vector Machine algorithms had the overall best ROC-AUC as well as balanced accuracy. The best performance for onset prediction achieved and ROC-AUC of 0.7699 for the onset prediction with SVM, and 0.6090 for the subtype prediction with RF.

**Table 3:** Classification Performance Summary

| *Onset Cohort* |  |  |  |  |  |
| --- | --- | --- | --- | --- | --- |
| Model | Balanced Accuracy | ROC-AUC | Precision | Recall | f1 Score |
| LR | 0.7020 | 0.7564 | 0.7629 | 0.7493 | 0.7560 |
| SVM | 0.7219 | 0.7699 | 0.7873 | 0.7403 | 0.7630 |
| LASSO | 0.6796 | 0.7298 | 0.7566 | 0.6866 | 0.7199 |
| ElasticNet | 0.6931 | 0.7430 | 0.7610 | 0.7224 | 0.7412 |
| RF | 0.7258 | 0.7659 | 0.7818 | 0.7701 | 0.7759 |
| GB | 0.6486 | 0.7629 | 0.7000 | 0.8149 | 0.7531 |
| XGB | 0.6544 | 0.7481 | 0.7071 | 0.8000 | 0.7507 |

| *Subtype Cohort* |  |  |  |  |  |
| --- | --- | --- | --- | --- | --- |
| Model | Balanced Accuracy | ROC-AUC | Precision | Recall | f1 Score |
| LR | 0.4639 | 0.5835 | 0.4661 | 0.4639 | 0.4636 |
| SVM | 0.5404 | 0.6037 | 0.5858 | 0.5404 | 0.5590 |
| LASSO | 0.3333 | 0.5552 | 0.1944 | 0.3333 | 0.2456 |
| ElasticNet | 0.4087 | 0.5464 | 0.2698 | 0.4087 | 0.2651 |
| RF | 0.4531 | 0.6090 | 0.4345 | 0.4531 | 0.4339 |
| GB | 0.2365 | 0.5124 | 0.1999 | 0.2366 | 0.2161 |
| XGB | 0.2987 | 0.5757 | 0.2592 | 0.2987 | 0.2750 |

### 2.4 Model Interpretation

The Random Forest model provides us with interpretable variable importance measures, calculated as the total reduction of sum of squared errors across all splits at which a given variable is used. Since RF was among the best performing algorithms for both onset prediction and subtype prediction tasks, we interpret our results by selecting the top 500 features with the highest importance of the RF models for the two tasks respectively. First, we split up any feature that contains more than one gene; some features contain more than one gene due to the fact that some genes overlap, and therefore some variants can be hosted by more than one gene. After this splitting, we are left with 520 important genes for the onset prediction task, and 547 important genes for the subtype prediction task. The overlap between these two gene sets are shown in Fig. 2. We then performed the Gene Set Enrichment Analysis https://www.gsea-msigdb.org/gsea/index.jsp) to these gene sets to see whether any biological pathways are enriched in these sets. The resulting pathways and their relative importance to the classification model are shown in Table. 4.

**Figure 2:**
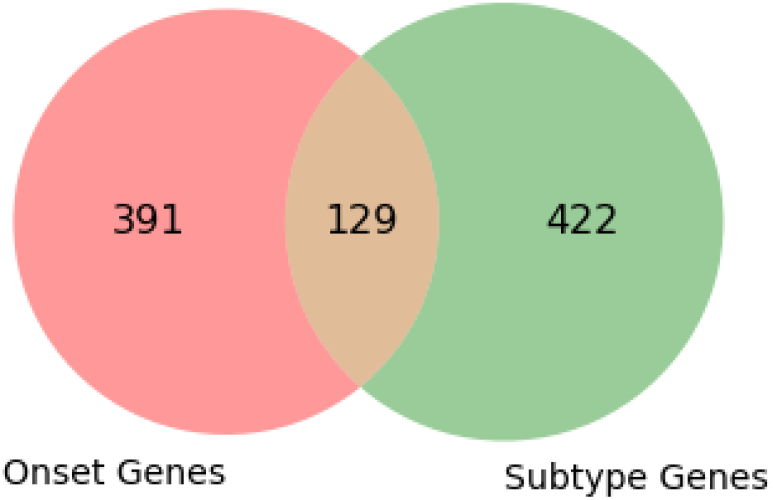
Venn diagram showing the overlap between the high-importance gene sets in PD onset prediction and subtype prediction.

**Table 4:** Gene set enrichment hits alongside their relative feature importance in percentage in the best-performing random forest classification model are shown. The FDR-Adjusted p-values for enrichment analysis are shown in parentheses.

| Enriched Pathways | Onset | Subtype |
| --- | --- | --- |
| Signaling by GPCR | 5.77 (3.4e-13) | 6.27 (3.4e-05) |
| Innate Immune System | 4.54 (4.3e-03) | 4.89 (2.4e-02) |
| Adaptive Immune System | 3.47 (1.2e-02) |  |
| Neutrophil degranulation | 2.85 (5.1e-03) | 2.58 (2.9e-02) |
| G alpha (s) signalling events | 2.72 (2.5e-13) | 4.72 (1.1e-07) |
| Olfactory Signaling Pathway | 2.31 (1.6e-14) | 4.45 (8.9e-10) |
| Metabolism of nucleotides | 1.01 (9.9e-04) |  |
| Diseases of glycosylation | 0.68 (4.0e-02) |  |
| Nucleotide salvage | 0.64 (2.0e-02) |  |
| Purine salvage | 0.59 (4.4e-02) |  |
| Diseases associated with O-glycosylation | 0.58 (2.1e-02) | 1.29 (3.4e-02) |
| Termination of O-glycan biosynthesis | 0.52 (1.7e-03) |  |
| HTFC/Tumoral calcinosis | 0.52 (4.2e-04) |  |
| TNPS/Polyagglutination syndrome | 0.52 (4.7e-04) |  |
| Dectin-2 family | 0.52 (2.8e-03) |  |
| Antigen Presentation | 0.41 (2.4e-02) |  |
| Nucleobase catabolism | 0.38 (1.1e-02) |  |
| Interferon Signaling |  | 2.75 (1.4e-04) |
| Interferon gamma signaling |  | 2.56 (1.5e-06) |
| Interferon alpha/beta signaling |  | 1.32 (8.0e-03) |
| Generation of second messenger molecules |  | 1.25 (2.4e-02) |
| PD-1 signaling |  | 1.25 (6.9e-03) |
| Endosomal/Vacuolar pathway |  | 0.73 (4.2e-02) |

Pathways with the highest total importance for both the onset prediction and subtype prediction tasks included immune-related pathways such as innate immune system and neutrophil degranulation pathways. Other common hits with high importance included signaling pathways such as olfactory signaling and GPCR signaling, as well as pathways related to diseases associated with O-glycosylation.

Pathways that were enriched only in the onset-related gene set included ones related to nucleotide metabolism such as nucleotide catabolism and purine salvage pathways, and other immune-related hits such as Dectin-2 family and antigen presentation pathways. The O-glycan synthesis pathway was also a unique hit for the Onset Genes. Pathways that were enriched only in the Subtype Genes included additional immune-related pathways, namely interferon-signaling and endosomal/vacuolar pathways.

## 3 Discussions

In this study, we trained machine learning models for two tasks, PD onset prediction and PD subtype prediction, using the number of deleterious variants per gene as the input features. The onset prediction task achieved a satisfactory performance of 0.77 ROC, and the subtype prediction task achieved a moderate performance of 0.60 macro-averaged

ROC, indicating that there seems to be some relationship between a person’s deleterious germline variants and their PD risk and prognosis. We interpreted our results by evaluating the variables of high importance in our top performing models.

For both of our tasks, Random Forest was one of the top-performing algorithms, with an ROC of 0.77 and 0.60 for the onset and subtype prediction, respectively. We derived high-importance gene sets for each task, and performed Gene Set Enrichment Analysis to obtain canonical biological pathways that were enriched in our high-importance gene sets.

One of the most notable patterns of our enrichment analysis result was the high number of immune-related pathway hits, some common between the Onset Genes and Subtype Genes, while others unique to one gene set. The adaptive immune system, innate immune system, and neutrophil degranulation were all pathways that were also defined in Bandres-Ciga *et al*. as pathways indicative of PD risk. Although some studies claim it is yet unknown whether the immune response is directly involved in the etiology of PD or simply a consequence of other PD processes such as *α*-synucleinopathy, [16] our results, together with others, suggests that the immune pathways likely play a role in the etiology of PD. Some ways in which the immune pathway could be related to PD etiology include an autoimmune response potentially responsible for dopaminergic neuronal death through neuroinflammation and degeneration. [17]

Interestingly, Signaling by GPCR pathway had a high contribution to both the onset and subtype classification tasks, which is again consistent with the results of Bandres-Ciga *et al*.. Most all genes that contributed to the Olfactory Signaling pathway as a hit were also a part of the GPCR pathway. This is a fascinating result, considering olfactory dysfucntion has long been known as a prodromal symptom of PD and have been used in its diagnosis. [18] In spite of being one of the earliest symptoms of PD, it is unknown whether olfactory decline is merely a symptom or is involved in disease etiology. One potential explanation behind olfactory decline is the accumulation of Lewy bodies in the olfactory bulb and its surrounding areas. Lewy pathology in the olfactory bulb is highly specific to PD, and its severity is known to correlate well with motor-symptoms. [19] It is hypothesized by some that the olfactory bulb is the induction site of Lewy pathology, which then spreads to the brain via the brainstem. [20] Taken together with our results, this highlights the possibility that certain germline mutations of genes related to olfactory function could have some causative relationship with disease course.

Hits in pathways related to protein glycation and glycosylation were also intriguing, considering the pathogenesis of PD. Protein glycation is widely known for its deleterious effects in aging [21]. Protein glycosylation affects protein structure and stability, and elevated levels of protein glycosylation have been associated with neurodegeneration for its effect on impairing clearance of dysfunctional proteins. [22] Several studies have established connections between PD and glycosylation as well as glycation. [23, 24]. Our results, taken together with these previous findings, imply that protein glycation and glycosylation could be involved in the development and progression of PD.

Another interesting aspect of the Gene Set Enrichment Analysis is that, although there was only 23-24% overlap between the two gene sets, the two gene sets shared many of the enriched pathways. It is possible that, although similar biologicaly pathways are involved in disease etiology, different genes, as components of pathways, contribute to separate aspects of the disease such as onset or subtype. Future studies might benefit from taking biological pathways into consideration when training predictive models.

Although our study provides a state-of-the-art PD prediction models based on germline variants, it still suffers from several limitations. The biggest limitation is the available sample size, particularly for the progression subtype prediction task. The sample size was limited to what was published in Zhang *et al*. due to the lack of standard definitions of progression subtypes in PD. The particular underrepresentation of Subtype II in our Subtype Cohort must also be taken into consideration when interpreting our models. However, the high performance of our Onset Prediction task is a promising implication that there indeed is some relationship behind PD disease course and a patient’s germline variants. Future studies would benefit from clearly defined progression subtype information in an integrated cohort study with a larger sample size such as AMP-PD. Another limitation of our current study is the lack of racial diversity in the AMP-PD cohorts. Many studies have shown that race plays a large role in PD risk, and therefore it is imperative to recruit a diverse cohort in future studies.

Overall, our study results provide insights that are of relevance to future biomedical and clinical research. Our high-importance gene sets call for further controlled laboratory experiments as well as clinical cohort studies to further confirm their role in the risk of PD development or defining its disease course. This could lead us not only to a better understanding of PD pathogenesis, but also to stronger clinical insights such as PD risk screening and patient-specific prognoses.

## 4 Conclusions

In this study, we developed predictive models for Parkinson’s Disease onset as well as progression subtypes using patients’ WGS and WES data, specifically using the counts of deleterious germline variants in each gene. We obtained gene sets that were top-importance in predicting the onset or subtype of PD, and interpreted the enriched pathways to draw connections to what is known about PD etiology. These gene sets provide us with promising candidates for future biomedical and clinical research.

## 5 Methods

### 5.1 Data processing and feature engineering

The files obtained from the AMP-PD and PPMI databases had been processed into Variant Call Format (VCF) files using the Genome Annotation Toolkit (GATK). [25] We first annotated the VCF files using the ANNOVAR toolkit, which provides attributes for each variant with information such as hosting gene, the variant’s functional effect, and its functional region. [26] We chose the well-established refGene database for this annotation. We then filtered out any variant with a low quality score (Phred-scaled base quality score<30, GQ<10) or low sequencing depth (DP<10). We only selected variants that are highly likely to be deleterious based on the variant’s functional effect, by selecting variants that are either frameshift insertions/deletions, splicing variants, stop-gain, or stop-loss variants. We did not perform any feature elimination based on known genomic features of PD to keep an unbiased approach. Finally, we generated a variant count matrix by counting the number of deleterious variants per gene in a given subject. Any feature (gene) in the resulting matrix that was either extremely common or rare (present in <5% or <95% of subjects) was eliminated as likely not contributing to disease pattern.

### 5.2 Model Training

We split our Onset Cohort and Subtype Cohort data into a training and held-out test set in a stratified manner to maintain the class composition in each set. We performed a 8:2 split on the Onset Cohort data and 9:1 split on the Subtype Cohort. We set the training set to be larger in the Subtype Cohort due to its small sample size. We trained seven different machine learning algorithms for our two tasks, respectively: logistic regression with L2 regularization (LR), LASSO, ElasticNet, Support Vector Machine (SVM), Random Forest Classifier (RF), Gradient Boosting Tree (GB), and XGBoost (XGB). We perform the a 5-fold cross-validation on the training set to tune the hyperparameters for our list of algorithms. Model training and evaluation were performed using a Python environment with Anaconda3, specifically with Scikit-learn 0.23.2 and the Python API of XGBoost 1.2.0. We account for class imbalance during model training by weighting the observations inversely proportional to their class count.

### 5.3 Model Evaluation and Interpretation

We evaluate our trained models by making predictions on our held-out test set. We calculate a number of evaluation metrics including balanced accuracy, area under the curve of receiver operating characteristic (ROC-AUC), precision, recall, and f1 score. Since there is class imbalance in our datasets, we macro-average our metrics across classes.

We then derive a set of 500 genes that had the highest importance in our best-performing models for the onset prediction task and the subtype prediction task, respectively. We ran Gene Set Enrichment Analysis to find out which biological pathways were enriched in our gene sets. We narrowed our search to canonical pathways characterized in the Reactome Pathway Database, and set a threshold of 0.05 for the FDR-adjusted p-value.

For each pathway that is significantly enriched in our gene sets, we evaluate the overall importance of the pathway by summing the importance scores of the genes that comprised that pathway.

The details of data processing and analysis have been made available at https://github.com/sayadennis/amppd as well as https://github.com/sayadennis/ppmi.

## Data Availability

All data used in this study are available from Accelerating Medicines Partnership: Parkinson's Disease (https://amp-pd.org/) and the Parkinson's Progression Marker's Initiative (https://www.ppmi-info.org/).

## References

[1] D. Hirtz, D. J. Thurman, K. Gwinn-Hardy, M. Mohamed, A. R. Chaudhuri, and R. Zalutsky. How common are the “common” neurologic disorders? Neurology, 68(5):326–337, Jan 2007.

[2] S. G. Reich and J. M. Savitt. Parkinson’s Disease. Med. Clin. North Am., 103(2):337–350, Mar 2019.

[3] U. K. Rinne, F. Bracco, C. Chouza, E. Dupont, O. Gershanik, J. F. Marti Masso, J. L. Montastruc, and C. D. Marsden. Early treatment of Parkinson’s disease with cabergoline delays the onset of motor complications. Results of a double-blind levodopa controlled trial. The PKDS009 Study Group. Drugs, 55 Suppl 1:23–30, 1998.

[4] S. Bandres-Ciga, S. Saez-Atienzar, J. J. Kim, M. B. Makarious, F. Faghri, M. Diez-Fairen, H. Iwaki, H. Leonard, J. Botia, M. Ryten, D. Hernandez, J. R. Gibbs, J. Ding, Z. Gan-Or, A. Noyce, L. Pihlstrom, A. Torkamani, A. R. Soltis, C. L. Dalgard, S. W. Scholz, B. J. Traynor, D. Ehrlich, C. R. Scherzer, M. Bookman, M. Cookson, C. Blauwendraat, M. A. Nalls, and A. B. Singleton. Large-scale pathway specific polygenic risk and transcriptomic community network analysis identifies novel functional pathways in Parkinson disease. Acta Neuropathol, 140(3):341–358, 09 2020.

[5] R. Chia, M. S. Sabir, S. Bandres-Ciga, S. Saez-Atienzar, R. H. Reynolds, E. Gustavsson, R. L. Walton, S. Ahmed, C. Viollet, J. Ding, M. B. Makarious, M. Diez-Fairen, M. K. Portley, Z. Shah, Y. Abramzon, D. G. Hernandez, C. Blauwendraat, D. J. Stone, J. Eicher, L. Parkkinen, O. Ansorge, L. Clark, L. S. Honig, K. Marder, A. Lemstra, P. St George-Hyslop, E. Londos, K. Morgan, T. Lashley, T. T. Warner, Z. Jaunmuktane, D. Galasko, I. Santana, P. J. Tienari, L. Myllykangas, M. Oinas, N. J. Cairns, J. C. Morris, G. M. Halliday, V. M. Van Deerlin, J. Q. Trojanowski, M. Grassano, A. Calvo, G. Mora, A. Canosa, G. Floris, R. C. Bohannan, F. Brett, Z. Gan-Or, J. T. Geiger, A. Moore, P. May, R. Krüger, D. S. Goldstein, G. Lopez, N. Tayebi, E. Sidransky, L. Norcliffe-Kaufmann, J. A. Palma, H. Kaufmann, V. G. Shakkottai, M. Perkins, K. L. Newell, T. Gasser, C. Schulte, F. Landi, E. Salvi, D. Cusi, E. Masliah, R. C. Kim, C. A. Caraway, E. S. Monuki, M. Brunetti, T. M. Dawson, L. S. Rosenthal, M. S. Albert, O. Pletnikova, J. C. Troncoso, M. E. Flanagan, Q. Mao, E. H. Bigio, E. Rodríguez-Rodríguez, J. Infante, C. Lage, I. González-Aramburu, P. Sanchez-Juan, B. Ghetti, J. Keith, S. E. Black, M. Masellis, E. Rogaeva, C. Duyckaerts, A. Brice, S. Lesage, G. Xiromerisiou, M. J. Barrett, B. S. Tilley, S. Gentleman, G. Logroscino, G. E. Serrano, T. G. Beach, I. G. McKeith, A. J. Thomas, J. Attems, C. M. Morris, L. Palmer, S. Love, C. Troakes, S. Al-Sarraj, A. K. Hodges, D. Aarsland, G. Klein, S. M. Kaiser, R. Woltjer, P. Pastor, L. M. Bekris, J. B. Leverenz, L. M. Besser, A. Kuzma, A. E. Renton, A. Goate, D. A. Bennett, C. R. Scherzer, H. R. Morris, R. Ferrari, D. Albani, S. Pickering-Brown, K. Faber, W. A. Kukull, E. Morenas-Rodriguez, A. Lleó, J. Fortea, D. Alcolea, J. Clarimon, M. A. Nalls, L. Ferrucci, S. M. Resnick, T. Tanaka, T. M. Foroud, N. R. Graff-Radford, Z. K. Wszolek, T. Ferman, B. F. Boeve, J. A. Hardy, E. J. Topol, A. Torkamani, A. B. Singleton, M. Ryten, D. W. Dickson, A. Chiò, O. A. Ross, J. R. Gibbs, C. L. Dalgard, B. J. Traynor, S. W. Scholz, A. R. Sotis, G. Sukumar, C. Alba, N. Lott, E. M. Martinez, M. Tuck, J. Singh, D. Bacikova, X. Zhang, D. N. Hupalo, A. Adeleye, M. D. Wilkerson, and H. B. Pollard. Genome sequencing analysis identifies new loci associated with Lewy body dementia and provides insights into its genetic architecture. Nat Genet, 53(3):294–303, 03 2021.

[6] C. Su, J. Tong, and F. Wang. Mining genetic and transcriptomic data using machine learning approaches in Parkinson’s disease. NPJ Parkinsons Dis, 6:24, 2020.

[7] B. Post, J. D. Speelman, and R. J. de Haan. Clinical heterogeneity in newly diagnosed Parkinson’s disease. J. Neurol., 255(5):716–722, May 2008.

[8] M. A. Thenganatt and J. Jankovic. Parkinson disease subtypes. JAMA Neurol, 71(4):499–504, Apr 2014.

[9] S. M. Fereshtehnejad and R. B. Postuma. Subtypes of Parkinson’s Disease: What Do They Tell Us About Disease Progression? Curr Neurol Neurosci Rep, 17(4):34, 04 2017.

[10] M. A. Thenganatt and J. Jankovic. Parkinson disease subtypes. JAMA Neurol, 71(4):499–504, Apr 2014.

[11] M. W. Libbrecht and W. S. Noble. Machine learning applications in genetics and genomics. Nat. Rev. Genet., 16(6):321–332, Jun 2015.

[12] J. A. Diao, I. S. Kohane, and A. K. Manrai. Biomedical informatics and machine learning for clinical genomics. Hum. Mol. Genet., 27(R1):R29–R34, 05 2018.

[13] X. Zhang, J. Chou, J. Liang, C. Xiao, Y. Zhao, H. Sarva, C. Henchcliffe, and F. Wang. Data-Driven Subtyping of Parkinson’s Disease Using Longitudinal Clinical Records: A Cohort Study. Sci Rep, 9(1):797, 01 2019.

[14] A. Landolfi, C. Ricciardi, L. Donisi, G. Cesarelli, J. Troisi, C. Vitale, P. Barone, and M. Amboni. Machine Learning Approaches in Parkinson’s Disease. Curr Med Chem, Jan 2021.

[15] K. S. Taylor, J. A. Cook, and C. E. Counsell. Heterogeneity in male to female risk for Parkinson’s disease. J Neurol Neurosurg Psychiatry, 78(8):905–906, Aug 2007.

[16] G. Gelders, V. Baekelandt, and A. Van der Perren. Linking Neuroinflammation and Neurodegeneration in Parkinson’s Disease. J Immunol Res, 2018:4784268, 2018.

[17] A. De Virgilio, A. Greco, G. Fabbrini, M. Inghilleri, M. I. Rizzo, A. Gallo, M. Conte, C. Rosato, M. Ciniglio Ap-piani, and M. de Vincentiis. Parkinson’s disease: Autoimmunity and neuroinflammation. Autoimmun Rev, 15(10):1005–1011, Oct 2016.

[18] M. E. Fullard, J. F. Morley, and J. E. Duda. Olfactory Dysfunction as an Early Biomarker in Parkinson’s Disease. Neurosci Bull, 33(5):515–525, Oct 2017.

[19] R. L. Doty. Olfactory dysfunction in Parkinson disease. Nat Rev Neurol, 8(6):329–339, May 2012.

[20] H. Braak and K. Del Tredici. Neuropathological Staging of Brain Pathology in Sporadic Parkinson’s disease: Separating the Wheat from the Chaff. J Parkinsons Dis, 7(1):S71–S85, 2017.

[21] I. Sadowska-Bartosz and G. Bartosz. Effect of glycation inhibitors on aging and age-related diseases. Mech. Ageing Dev., 160:1–18, 12 2016.

[22] W. Y. Wani, J. C. Chatham, V. Darley-Usmar, L. L. McMahon, and J. Zhang. O-GlcNAcylation and neurodegeneration. Brain Res. Bull., 133:80–87, Jul 2017.

[23] L. Ma, J. Song, X. Sun, W. Ding, K. Fan, M. Qi, Y. Xu, and W. Zhang. Role of microtubule-associated protein 6 glycosylated with Gal-(Î^2^-1,3)-GalNAc in Parkinson’s disease. Aging (Albany NY), 11(13):4597–4610, 07 2019.

[24] J. P. Trezzi, S. Galozzi, C. Jaeger, K. Barkovits, K. Brockmann, W. Maetzler, D. Berg, K. Marcus, F. Betsou, K. Hiller, and B. Mollenhauer. Distinct metabolomic signature in cerebrospinal fluid in early parkinson’s disease. Mov. Disord., 32(10):1401–1408, Oct 2017.

[25] A. McKenna, M. Hanna, E. Banks, A. Sivachenko, K. Cibulskis, A. Kernytsky, K. Garimella, D. Altshuler, S. Gabriel, M. Daly, and M. A. DePristo. The Genome Analysis Toolkit: a MapReduce framework for analyzing next-generation DNA sequencing data. Genome Res., 20(9):1297–1303, Sep 2010.

[26] K. Wang, M. Li, and H. Hakonarson. ANNOVAR: functional annotation of genetic variants from high-throughput sequencing data. Nucleic Acids Res., 38(16):e164, Sep 2010.

